# Healthcare-Delivered Lifestyle Recommendations and Their Association with Health Behaviors in Iran

**DOI:** 10.64898/2026.03.11.26348114

**Authors:** Hanye Sohrabi, Mina Mirzad, Ali Golestani, Sina Azadnajafabad, Naser Ahmadi, Arian Afzalian, Nazila Rezaei, Mohammad-Mahdi Rashidi, Erfan Ghasemi, Negar Rezaei, Moein Yoosefi, Ameneh Kazemi, Shirin Djalalinia, Yosef Farzi, Rosa Haghshenas, Maryam Nasserinejad, Elmira Foroutan Mehr, Sahar Mohammadi Fateh, Farshad Farzadfar

**Author notes:** Corresponding author: (FF). These authors contributed equally to this work (Hanye Sohrabi, Mina Mirzad, Ali Golestani).

## Abstract

**Background:** Non-communicable diseases (NCDs) are the leading cause of mortality in Iran, driven largely by modifiable lifestyle risk factors. Healthcare providers play a pivotal role in delivering preventive lifestyle recommendations, yet the extent and effectiveness of these efforts remain unclear. This study aimed to assess the distribution of lifestyle counseling across sociodemographic and clinical subgroups and its association with health behavior status in Iran using data from the 2021 WHO STEPS survey.

**Methods:** We conducted a cross-sectional analysis of 27,704 included adults participating in the nationally representative 2021 STEPS survey in Iran. Data on delivery of ten specific lifestyle recommendations within the past 12 months, covering diet, physical activity, weight management, and tobacco non-initiation/cessation, were collected. Associations between recommendation delivery and health behaviors were analyzed using logistic regression models.

**Results:** Only 33.4% (95% Confidence Interval (CI): 32.8-34.0) of participants received all ten lifestyle recommendations, while 10.7% (10.2-11.1) received no recommendations. Recommendations were more frequently delivered to females, rural residents, and individuals with multiple comorbidities, indicating a risk-based response by healthcare providers. Notably, tobacco non-initiation/cessation advice was not more commonly offered to patients with multiple chronic conditions. Delivery of lifestyle counseling was associated with positive behavior status: individuals who received weight loss/ maintaining normal weight and physical activity advice were more likely to engage in sufficient physical activity (adjusted odds ratio 1.21(1.13-1.29)), and among current smokers, receiving tobacco cessation recommendation was associated with higher tobacco quitting attempt (aOR: 1.83(1.49-2.24)). A dose-response relationship was observed between the number of nutritional recommendations received and better diet quality (aOR for 6 vs. ≤2 recommendations: 1.32 (1.24-1.41)). Geographical disparities were evident, with eastern provinces receiving the least comprehensive counseling.

**Conclusion:** Delivery of lifestyle recommendations by healthcare providers in Iran shows variation by sociodemographic and clinical factors and is positively associated with behavior status. These findings highlight the effectiveness of provider-delivered lifestyle counseling and the need for more consistent, equitable, and targeted delivery, particularly for high-risk individuals and underserved regions, to strengthen national NCD prevention efforts.

## Introduction

Non-communicable diseases (NCDs) represent a major global health challenge, responsible for an estimated 43 million deaths worldwide in 2021, according to the World Health Organization (WHO). Four major NCDs, cardiovascular diseases, cancers, chronic respiratory diseases, and diabetes, account for nearly 80% of all NCD-related deaths globally, with 82% of these occurring in low- and middle-income countries (LMICs) [1]. These conditions are closely linked to modifiable lifestyle risk factors, including poor nutrition, physical inactivity, excess body weight, tobacco use, and harmful alcohol consumption, which necessitate targeted and evidence-based intervention programs [2].

In Iran, a middle-income country, NCDs account for approximately 86% of all deaths, placing a substantial burden on the healthcare system and impeding broader socio-economic development [3]. Most of the leading NCD risk factors in Iran are either behavioral, such as smoking and dietary habits, or metabolic, including hypertension, obesity, hyperglycemia, and dyslipidemia [4]. Reducing the population-level burden of these risk factors is the key to diminishing the rising impact of NCDs.

Healthcare providers across the continuum of care, from primary care physicians to specialists and interprofessional teams, are integral to health promotion and disease prevention by delivering lifestyle recommendations as part of routine clinical practice.[5,6] While preventive counseling has been emphasized in international guidelines for cardiovascular disease, diabetes, chronic respiratory disease, and cancer[7–11], there remains limited evidence on how effectively such recommendations are delivered in practice, particularly in low- and middle-income countries. In Iran, several national strategies have been developed to operationalize this role within the health care system. Aligned with the WHO Global Action Plan for the Prevention and Control of NCDs[12], the Iranian Non-Communicable Diseases Committee (INCDC) was established through multisectoral collaboration and launched a national action plan in 2015. [13,14]Despite these policy-level efforts, little is known about how lifestyle recommendations are distributed across the population and whether their receipt translates into measurable behavior change. The WHO STEPwise approach to Surveillance (STEPS) provides a standardized method for collecting nationally representative data on NCDs and their risk factors [15]. Using data from the 2021 STEPS survey [16], this study aims to examine the extent to which lifestyle recommendations are delivered by healthcare providers during routine medical encounters, and to evaluate whether receiving such advice is associated with positive behavior change among Iranian adults. By addressing this knowledge gap, the study offers important insights into the effectiveness of current NCD prevention strategies and identifies areas for improvement within Iran’s healthcare system.

## Methods

### Study Design and Participants

This research utilized the data from the STEPS 2021 study [17]. The STEPS 2021 dataset was accessed on October 26, 2024 (08/26/2024). The Iranian STEPS 2021 survey was designed to provide data on NCDs risk factors through a three-step methodology: (1) questionnaire-based interviews, (2) physical measurements, and (3) laboratory assessments. A systemic cluster random sampling strategy was administered across all 31 provinces, based on population size and relative weighting to ensure representativeness at both national and subnational levels. Of the 28,821 estimated sample size, 27,874 individuals completed the first step, 27,745 underwent physical measurements, and 18,119 participants aged 25 years or older provided biological samples for laboratory testing. Only Iranian adults aged 18 years and older were eligible, and the presence of any mental or physical conditions that prevented participation in the study steps and pregnancy were considered exclusion criteria. For the purpose of this study, we conducted a secondary analysis of the STEPS 2021 dataset and excluded 170 individuals due to missing responses for the lifestyle recommendation questions.

### Data Collection

In the first steps of the STEPS survey, information was gathered through a standardized questionnaire that addressed a wide range of topics, including demographic characteristics, dietary habits, medical history, physical activity, lifestyle recommendation, tobacco and alcohol consumption, and household assets. The second step involved collecting physical measurements including height, weight, waist, hip circumference, and blood pressure. Height was measured with a standard stadiometer, with participants standing straight against a wall, ensuring contact at the heels, hips, and back of the head. Body weight was measured using a calibrated Inofit digital scale, with participants dressed in light clothing and barefoot. Prior to recording blood pressure, participants were seated and rested for 15 minutes. Blood pressure was then measured three times at three-minute intervals using calibrated Beurer sphygmomanometer, with the final value being the average of the second and third readings. The third step involved serum and spot urine samples. Biological samples were preserved and transported under cold chain conditions to a central laboratory for further analysis.

### Definition of Variables

The main outcome of this study was the receipt of ten lifestyle recommendations from physicians or other healthcare providers within the past 12 months. Participants self-reported whether they had received recommendations on the following: tobacco non-initiation/cessation, reducing salt intake, daily consumption of fruits and vegetables, reducing red meat and processed foods, lowering fat intake, increasing fish consumption, consumption of whole grain bread and complex carbohydrates, reducing sugar intake, exercising, and losing weight/maintaining normal weight. Based on the number of recommendations received, participants were categorized into four groups: those who received all ten recommendations, those who received six to nine, those who received one to five, and those who did not receive any recommendation.

This study assessed multiple demographics, behavioral, and clinical variables using the 2021 WHO STEPS survey questionnaire and associated measurements. Participants were categorized into three age groups: 18-39 years, 40-59 years, and 60 years or older. Area of residence was classified as either urban or rural, and marital status was grouped into married, divorced or widowed, and never married. Educational attainment was categorized into three groups: illiterate or 1-6 years of schooling, 7-11 years, and 12 or more years of education. Employment status was recorded as employed, retired, unemployed, or unpaid. Socioeconomic status was measured using the wealth index, which was calculated from 36 household asset-related questions using principal component analysis (PCA). The first principal component, which captured the greatest variation among variables, was selected as the summary measure and categorized into five quintiles, ranging from the poorest (first quintile) to the wealthiest (fifth quintile) segments of the population, in accordance with the STEPS 2021 protocol [16]. Health insurance coverage was recorded as basic only, basic plus complementary, or no insurance.

The presence of chronic conditions, including diabetes, hypertension, and hypercholesterolemia was based on self-reported ever diagnoses by healthcare providers. This approach allowed us to focus on individuals aware of their diagnosis. Other comorbidities were similarly defined based on self-report. Participants were considered to have cardiovascular disease (CVD) if they reported ever being told by a healthcare worker that they had a heart attack, angina, or stroke. For chronic respiratory disease, participants were classified as having the condition if they have ever been diagnosed with asthma or chronic obstructive pulmonary disease (COPD), based on self-report of long-term respiratory symptoms and physician diagnosis. Cancer status was defined by the self-report of diagnosis of any types of cancer in the past 12 months. The number of comorbidities was calculated as the count of the following conditions: hypertension, diabetes, hypercholesterolemia, CVDs, chronic respiratory disease, and cancer. We then categorized participants into three groups: 0, 1, or ≥2 comorbidities.

BMI was calculated as weight in kilograms divided by height in meters squared (kg/m²). Participants were classified as underweight or normal weight (BMI < 25 kg/m²), overweight (BMI 25 to <30 kg/m²), or obese (≥30 kg/m²). Physical activity was assessed using the Global Physical Activity Questionnaire (GPAQ). Based on total weekly metabolic equivalent of task (MET), individuals with <600 MET-minutes/week were considered to have insufficient physical activity, per WHO guidelines [18].

Regarding tobacco and alcohol consumption, current tobacco use was defined as any occasional or daily use of tobacco products during the past year. Individuals who reported ever using tobacco but had not used it in the past year were classified as past smokers. Passive smokers were identified as those who reported exposure to secondhand smoke in home or workplace. Participants who reported no history of tobacco use and no exposure to secondhand smoke were considered never smokers. For alcohol consumption, individuals were classified as current drinkers if they had consumed alcohol within the past 12 months, as past drinkers if they had previously consumed alcohol but not within the past year, and as never drinkers if they had never consumed alcohol.

Nutritional practices were evaluated using six key dietary components: intake of fruits, vegetables, red meat, fish, whole grains, and fats. Each component was scored from 0 to 1 based on participants’ responses on the serving/frequency of consumption, reflecting alignment with dietary guidelines and expert recommendations (S1 Table). These scores were then summed to calculate a composite nutrition score with possible values ranging from 0 to 6, where higher scores indicate better dietary quality. Salt intake was assessed, using an indirect estimate of daily salt consumption derived from urinary sodium concentration measured in spot urine samples collected during the laboratory phase of the STEPS 2021 survey. Salt intake was classified as either appropriate or inappropriate based on whether it fell below or above the population median value of 9.60 grams per day.

### Statistical Analysis

Descriptive statistics were used to summarize the characteristics of the study population. To achieve both national and provincial representativeness, a four-step weighting procedure was applied: (1) adjustment for overall non-response, (2) adjusting for non-response specific to each survey step, (3) aligning the sample distribution with provincial population structures by age, sex, and area of residence, and (4) integrating all adjustments into a final composite weight [19]. Categorical variables were reported as weighted prevalence with 95% confidence intervals (CIs). To assess the associations between receiving different lifestyle modification recommendations and various outcomes, logistic regression was used for binary response variables, and ordinal logistic regression was employed for response variables with more than two categories. For each response variable, three models were constructed: (1) a crude model, (2) an age- and sex-adjusted model, and (3) a multivariate model adjusted for covariates selected through a stepwise approach with a p-value threshold of <0.2. Statistical significance was determined at a p-value threshold of <0.05. All analyses were conducted using R 4.4.1, and the ‘survey’ package was used to account for the complex survey design.

### Ethics Approval and Consent to Participate

Ethical approval for this study was obtained from the Research Ethics Committee of the Endocrine & Metabolism Research Institute, Tehran University of Medical Sciences (Approval ID: IR.TUMS.EMRI.REC.1403.056; Approval date: 05 August 2024). The analysis was conducted using anonymized data from the 2021 WHO STEPS survey. The STEPS survey itself received ethical approval from the relevant national ethics committee, and all participants provided informed consent prior to participation. No identifiable personal information was accessed by the authors.

## Results

### Population Characteristics

Among 27,704 participants, 44.7% were males and 75.0% lived in urban areas. The age distribution was 39.4% (18-39 years), 39.2% (40-59 years), and 21.4% (≥60 years). Most were married (76.6%) and 43.6% had ≥12 years of education. Employment status showed 37.0% were employed and 48.5% unpaid workers. Basic insurance covered 61.9% of participants. Self-reported diabetes, hypertension, and hypercholesterolemia were 10.5%, 21.8%, and 17.8%, respectively. Obesity was 25.0%, while 50.5% had insufficient physical activity. Current tobacco and alcohol use were reported in 14.0% and 3.8%, respectively (Table 1).

**Table 1.**
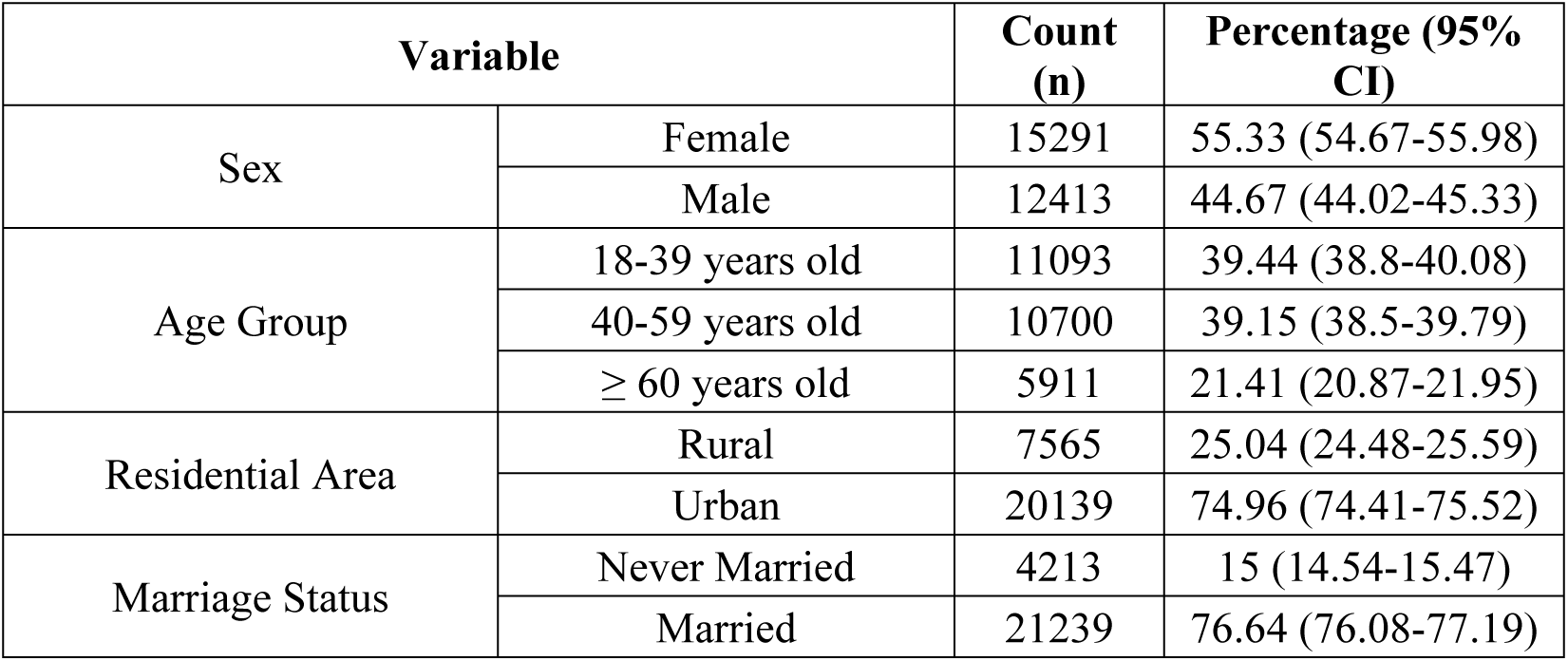
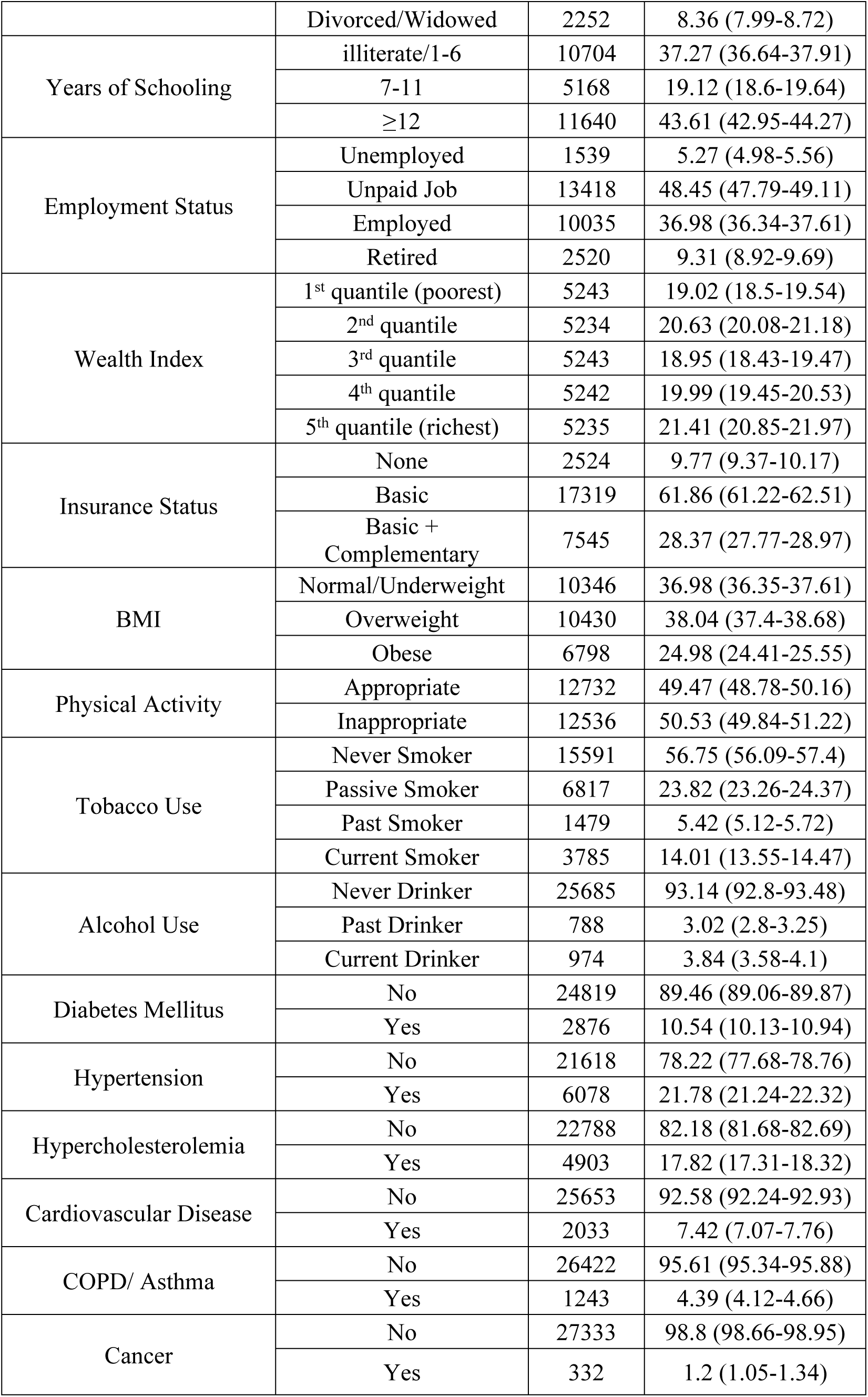
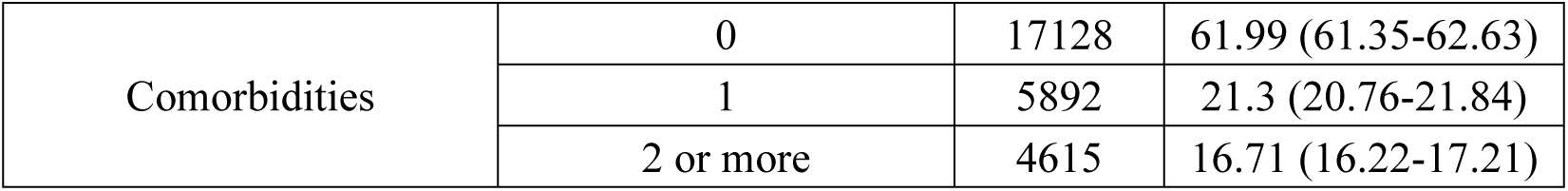
Baseline Characteristics of the Study Population Including Sociodemographic, Behavioral, and Clinical Variables.

### Distribution of Lifestyle Recommendations Across Sociodemographic Groups

Table 2 presents the prevalence of lifestyle recommendations across different subgroups. Totally, 33.4% (95%CI: 32.8-34.0) of participants received all of the recommendations, 35.8% (35.2-36.5) received six to nine recommendations, 20.1% (19.6-20.7) received less than six recommendations, and 10.65% (10.2-11.1) received no recommendations. A provincial comparison revealed that the proportion of individuals receiving all ten lifestyle recommendations was lowest in the eastern provinces of Iran and highest in the northwest. An approximately inverse pattern was observed for the proportion of individuals who received no recommendation (Fig 1). Among nutritional recommendations, the most frequently delivered nutritional recommendation was to increase daily fruit and vegetable consumption, received by 77.0% (95% CI: 76.4-77.6) of participants, followed by advice to reduce fat intake (72.5%, 71.9-73.0), decrease salt consumption (71.0%, 70.4-71.6), reduce sugar intake (71.3%, 70.7-71.9), consume less red meat and processed foods (67.0%, 66.4-67.6), increase fish intake (69.0%, 68.3–69.6), and increase whole grain bread and complex carbohydrates (65.0%, 64.3-65.6). Among behavioral recommendations, advice to increase physical activity was the most commonly delivered (72.3%, 71.7-72.9), followed by weight loss/maintaining normal weight recommendation (69.1%, 68.5-67.0). Tobacco non-initiation/cessation advice was the least commonly reported, received by 49.8% of participants (49.2-50.5).

**Figure 1.**
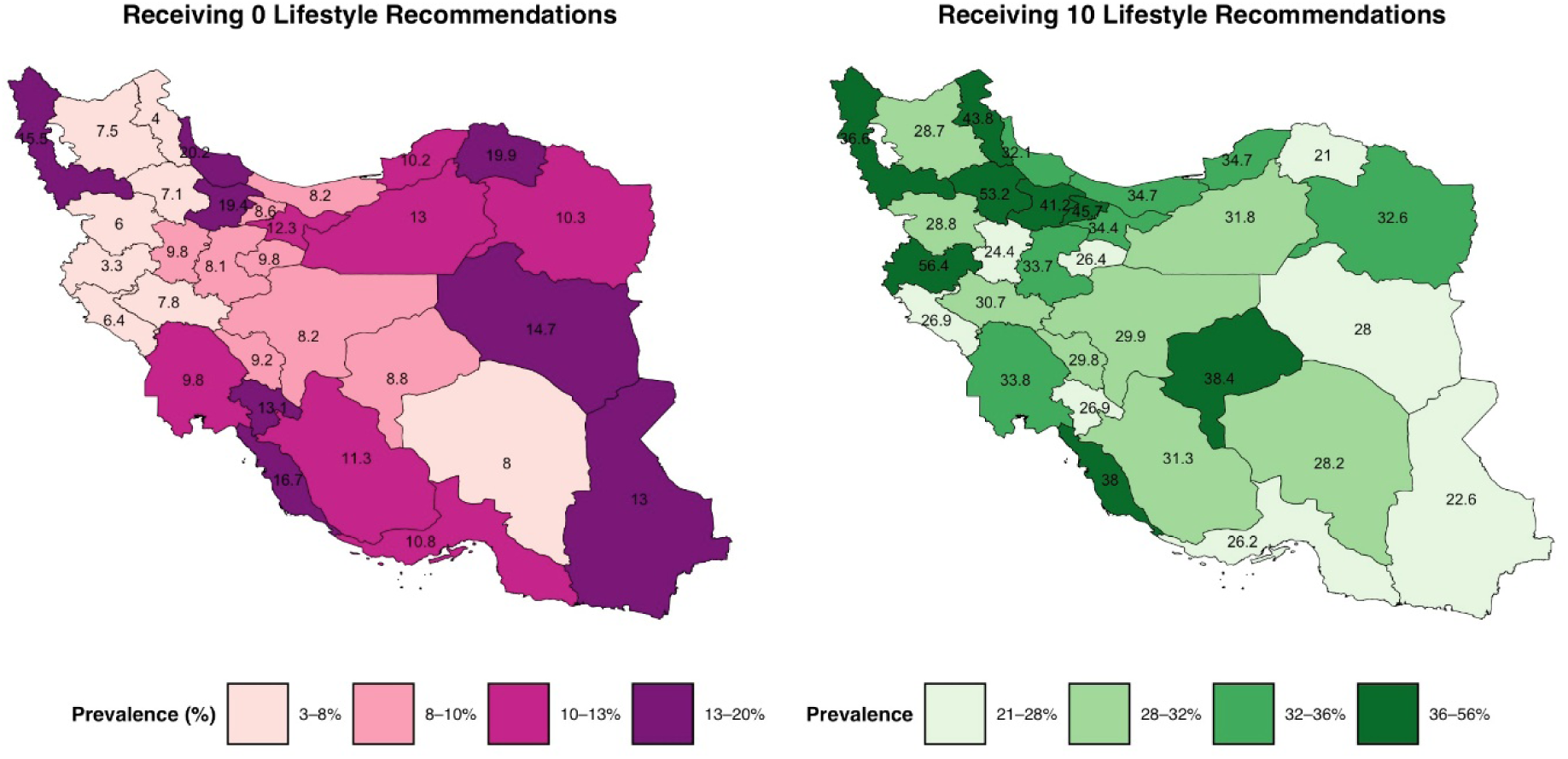
Weighted prevalence of individuals who received all ten recommendations and those who received none by province.

**Table 2.**
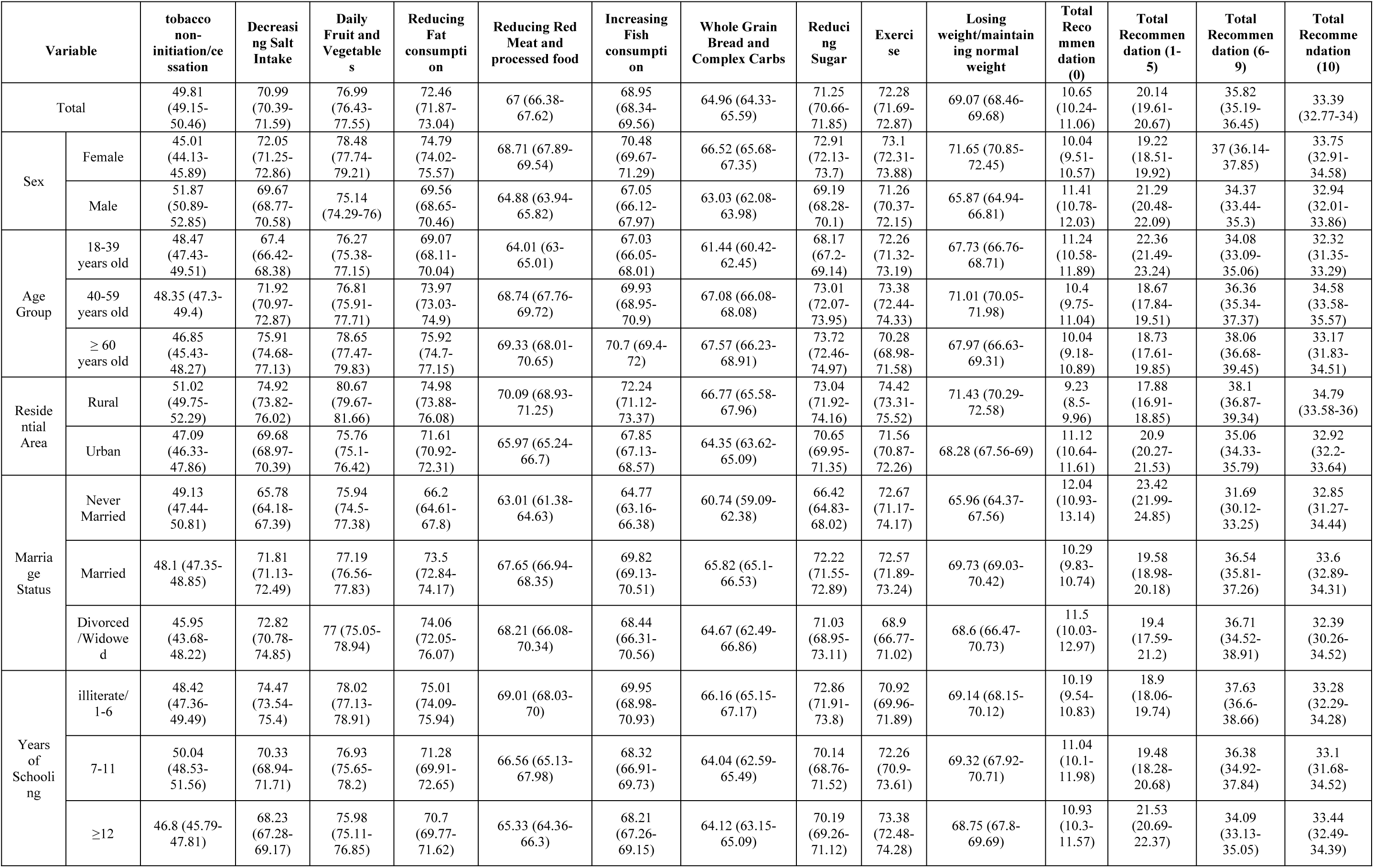
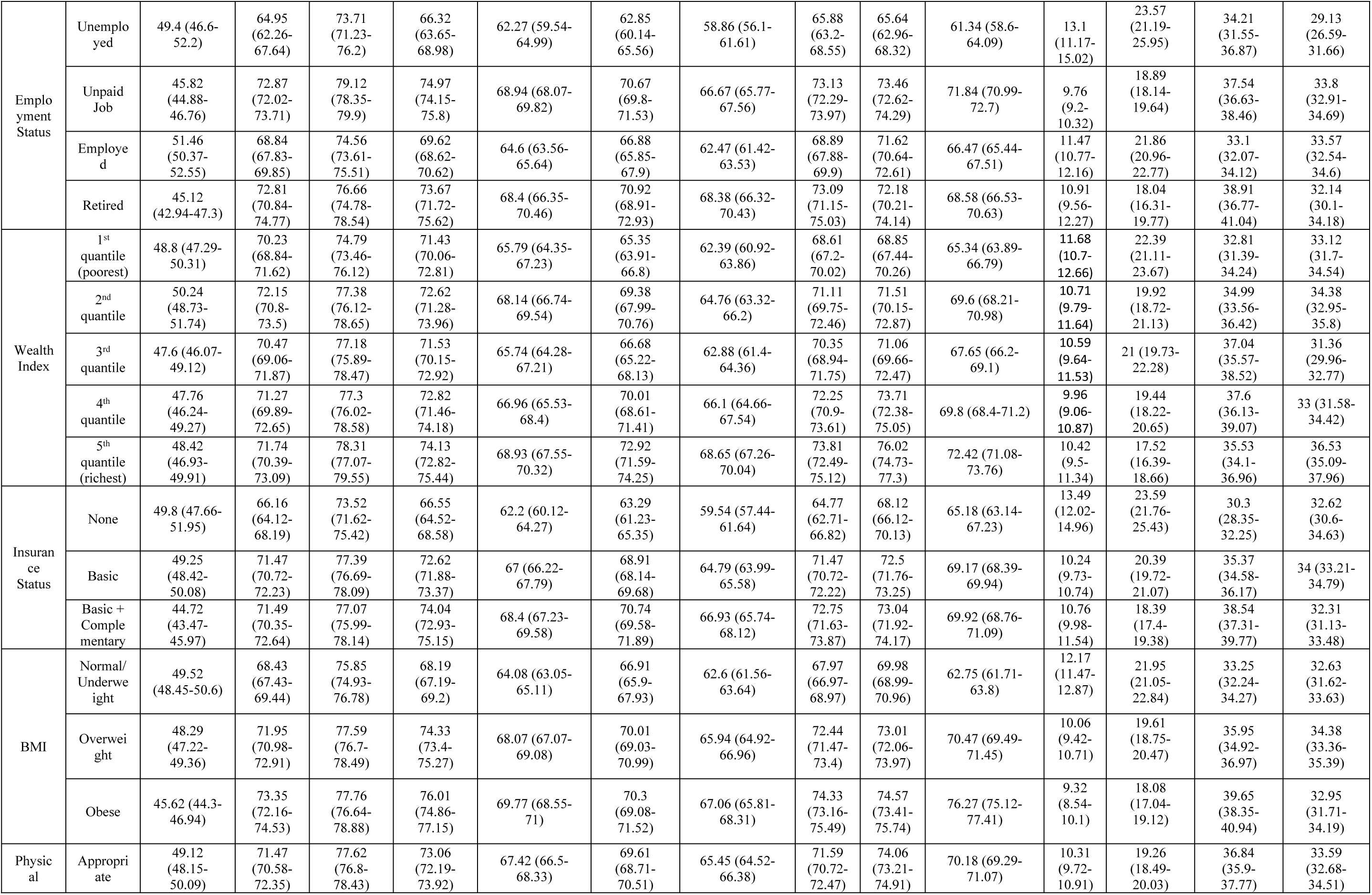
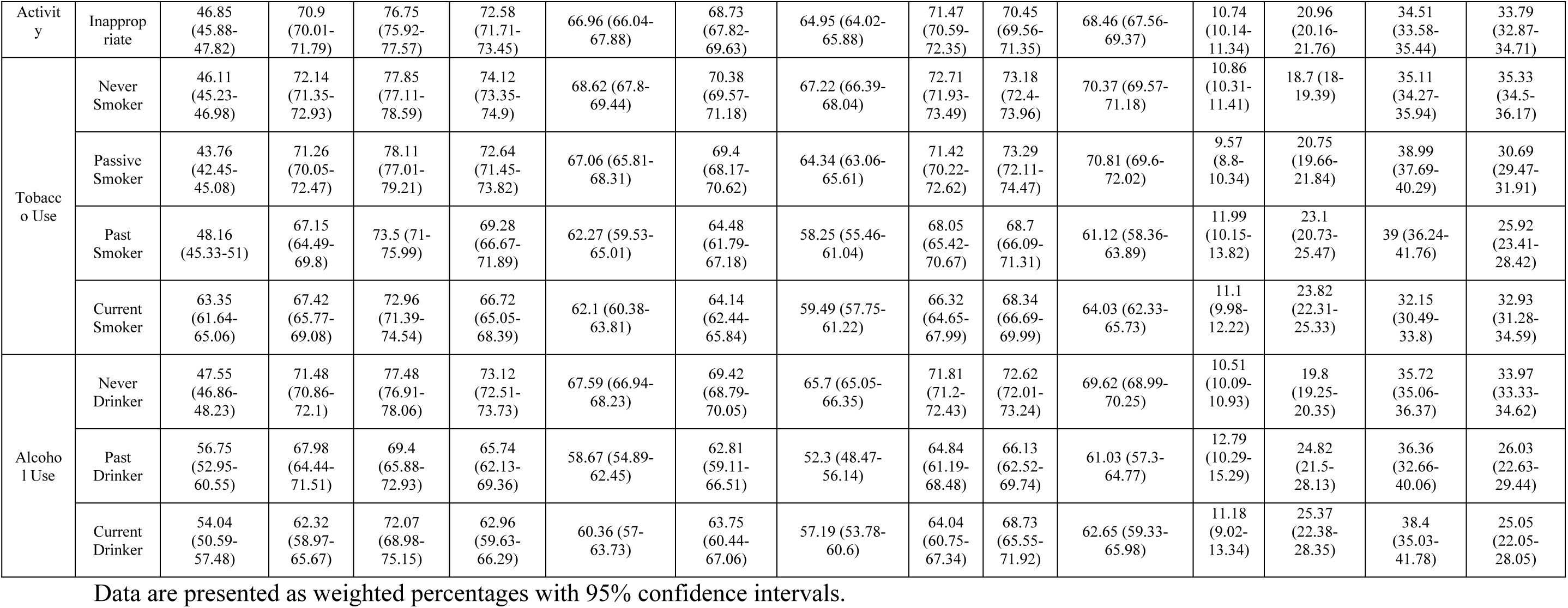
Prevalence of Lifestyle Recommendations Received by Sociodemographic and Behavioral Characteristics.

Most lifestyle recommendations were delivered more frequently to female participants compared to males, with the exception of tobacco non-initiation/cessation, which was more often recommended to males (51.9% [95%CI: 50.9-52.9] among males vs. 45.0% [44.13-45.89] in females). For most lifestyle recommendations, the sex-specific difference in delivery of lifestyle recommendations were unremarkable, with prevalence in females generally ranging between 66% and 75%, and in males between 63% and 71%, across nutritional and behavioral advice categories.

Participants living in rural areas received lifestyle recommendations more often than those in urban settings, for advice on dietary improvement, physical activity, weight management, and non-initiation/ cessation tobacco. The dietary recommendation delivery prevalence ranged from 66.8% to 80.7% in rural areas, and from 64.4% to 75.8% in urban areas. On the other hand, the behavioral recommendation prevalence ranged from 51.0% (tobacco non-initiation/cessation) to 74.4% (exercise) in rural areas, and 47.1% (tobacco non-initiation/cessation) to 71.6% (exercise) in urban areas. Other sociodemographic factors, such as age, marital status, education level, and employment status—showed recommendation patterns similar to the overall sample, with no major deviations in prevalence observed.

### Distribution of Lifestyle Recommendations Across Behavioral Risk Factors

Across BMI categories, several lifestyle recommendations were more frequently provided to individuals with higher BMI, showing a consistent increase from the normal-weight group to the overweight and obese groups. These included advice to reduce salt intake (from 68.4% [95% CI: 67.4-69.4] in the normal-weight group to 73.4% [72.2-74.5] in the obese group), lower fat consumption (from 68.2% [67.2-69.2] to 76.0% [74.7-77.2]), decrease red and processed meat intake (from 64.1% [63.1-65.1] to 69.8% [68.6-71.0]), increase fish consumption (from 66.9% [65.9-67.9] to 70.3% [69.1-71.5]), increase whole grain intake (from 62.6% [61.6-63.6] to 67.1% [65.8-68.3]), and reduce sugar intake (from 68.0% [67.0-69.0] to 74.3% [73.2-75.5]). Recommendations to increase physical activity also followed this pattern (from 70.0% [69.0-71.0] to 74.6% [73.4-75.6]). No statistically significant differences were observed between the overweight and obese groups for these recommendations. However, a notable exception was the recommendation to lose weight/maintain normal weight, which increased from 62.8% [61.7-63.8] in the normal-weight group to 70.5% [69.5-71.5] in the overweight group and further to 76.3% [75.1-77.4] in the obese group (Table 2).

Additionally, the only recommendation that was significantly more common among those with insufficient physical activity was the advice to exercise (74.1%, 95% CI: 73.2-75.0). Current smokers were significantly more advised to quit smoking (63.4%, 61.6-65.1), yet they received other health recommendations less than those who had never smoked. Moreover, tobacco non-initiation/cessation was generally less delivered to all subgroups compared to other recommendations. In terms of alcohol use, current and past consumers of alcohol only received the recommendation of non-initiation/cessation tobacco (54.0 [95%CI: 50.6-57.5] in current consumers and 56.8% [53.0-61.0] in past consumers) more than those who never consumed alcohol (47.6%, 46.9-48.2). Other recommendations were more often delivered to participants who never consumed alcohol (Table 2).

### Distribution of Lifestyle Recommendations Across Comorbidity Groups

When comparing the frequency of recommendation delivery across different comorbid groups, certain patterns emerged. In general, the prevalence of receiving lifestyle recommendations increased with the number of comorbidities, except for smoking non-initiation/cessation, which showed an insignificant trend (Table 3). Overall, participants with diabetes, hypertension, hypercholesterolemia, or CVD received almost all recommendations more frequently than participants without these conditions, except for smoking non-initiation/cessation, which showed no difference. Individuals with cancer were the most likely to receive recommendations to reduce salt intake (81.5, 95%CI: 76.9-86.2), increase fruit and vegetable consumption (83.0,78.5-87.5), and reduce fat intake (82.2, 77.6-86.8). Recommendations to reduce sugar consumption were given most frequently to individuals with diabetes (82.19, 80.63-83.75). In contrast, recommendations to reduce red meat consumption, increase fish intake, engage in more physical activity, and lose weight/maintain normal weight were distributed relatively evenly across different comorbid groups.

**Table 3.** Prevalence of Lifestyle Modification Recommendations by Chronic Disease Status in Adults.

| Variable |  | tobacco non-initiation /cessation | Decreasing Salt Intake | Daily Fruit and Vegetable | Reducing Fat consumption | Reducing Red Meat and processed food | Increasing Fish consumption | Whole Grain Bread and Complex Carbs | Reducing Sugar | Exercise | Losing weight/maintaining normal weight | Total Recommendation (0) | Total Recommendation (1-5) | Total Recommendation (6-9) | Total Recommendation (10) |
| --- | --- | --- | --- | --- | --- | --- | --- | --- | --- | --- | --- | --- | --- | --- | --- |
| Diabetes Mellitus | No | 48.26 (47.57-48.96) | 70.25 (69.61-70.89) | 76.64 (76.05-77.23) | 71.71 (71.08-72.34) | 66.32 (65.66-66.98) | 68.42 (67.77-69.06) | 64.17 (63.5-64.83) | 69.96 (69.32-70.6) | 71.64 (71.01-72.27) | 68.27 (67.62-68.92) | 11.04 (10.6-11.48) | 20.66 (20.1-21.23) | 35 (34.33-35.66) | 33.3 (32.65-33.95) |
|  | Yes | 46.42 (44.39-48.45) | 77.26 (75.54-78.97) | 79.96 (78.31-81.6) | 78.77 (77.09-80.45) | 72.74 (70.92-74.57) | 73.39 (71.59-75.2) | 71.65 (69.81-73.49) | 82.19 (80.63-83.75) | 77.65 (75.95-79.35) | 75.85 (74.1-77.6) | 7.36 (6.3-8.43) | 15.7 (14.21-17.2) | 42.92 (40.9-44.94) | 34.01 (32.09-35.94) |
| Hypertension | No | 48.47 (47.73-49.22) | 69.04 (68.35-69.73) | 76.03 (75.39-76.67) | 70.85 (70.17-71.52) | 65.93 (65.22-66.63) | 68.02 (67.32-68.71) | 63.92 (63.2-64.63) | 69.9 (69.22-70.58) | 71.59 (70.92-72.27) | 67.91 (67.21-68.61) | 11.33 (10.85-11.8) | 20.85 (20.24-21.46) | 34.38 (33.67-35.08) | 33.45 (32.75-34.15) |
|  | Yes | 46.62 (45.22-48.02) | 77.98 (76.81-79.15) | 80.41 (79.28-81.53) | 78.21 (77.05-79.38) | 70.79 (69.5-72.08) | 72.27 (71-73.53) | 68.63 (67.33-69.94) | 76.09 (74.88-77.29) | 74.68 (73.45-75.9) | 73.21 (71.96-74.45) | 8.24 (7.47-9.01) | 17.61 (16.53-18.69) | 41.04 (39.66-42.43) | 33.11 (31.79-34.43) |
| Hypercholesterolemia | No | 48.67 (47.94-49.39) | 70.2 (69.53-70.86) | 76.6 (75.98-77.22) | 70.98 (70.32-71.64) | 66.06 (65.37-66.74) | 68.19 (67.51-68.87) | 64.19 (63.5-64.89) | 70.14 (69.48-70.81) | 71.64 (70.98-72.29) | 68.14 (67.46-68.82) | 11.26 (10.8-11.72) | 20.56 (19.97-21.15) | 34.54 (33.85-35.23) | 33.64 (32.96-34.32) |
|  | Yes | 45.29 (43.73-46.85) | 74.63 (73.26-76) | 78.76 (77.46-80.05) | 79.25 (77.97-80.53) | 71.35 (69.92-72.77) | 72.41 (71.01-73.82) | 68.48 (67.02-69.94) | 76.34 (75.01-77.67) | 75.19 (73.84-76.54) | 73.35 (71.96-74.74) | 7.84 (7.01-8.68) | 18.25 (17.02-19.48) | 41.74 (40.2-43.29) | 32.17 (30.71-33.63) |
| Cardiovascular Disease | No | 48.02 (47.34-48.71) | 70.58 (69.96-71.21) | 76.78 (76.2-77.36) | 72.07 (71.45-72.68) | 66.67 (66.03-67.32) | 68.75 (68.12-69.39) | 64.69 (64.03-65.34) | 70.98 (70.36-71.6) | 72.27 (71.66-72.89) | 68.92 (68.28-69.55) | 10.79 (10.36-11.21) | 20.32 (19.76-20.87) | 35.44 (34.78-36.09) | 33.46 (32.82-34.1) |
|  | Yes | 48.69 (46.26-51.12) | 76.22 (74.16-78.28) | 79.91 (77.96-81.86) | 77.48 (75.43-79.52) | 71.12 (68.91-73.33) | 71.51 (69.31-73.71) | 68.5 (66.24-70.76) | 74.74 (72.62-76.86) | 72.52 (70.36-74.69) | 71.1 (68.9-73.3) | 8.86 (7.48-10.24) | 17.82 (15.95-19.69) | 40.89 (38.5-43.28) | 32.43 (30.16-34.7) |
| COPD/ Asthma | No | 48.11 (47.44-48.79) | 70.93 (70.31-71.54) | 76.96 (76.39-77.53) | 72.32 (71.72-72.93) | 66.85 (66.22-67.49) | 68.83 (68.21-69.46) | 64.82 (64.18-65.47) | 71.17 (70.56-71.78) | 72.2 (71.6-72.81) | 68.91 (68.28-69.53) | 10.74 (10.32-11.16) | 20.15 (19.61-20.7) | 35.62 (34.97-36.26) | 33.49 (32.85-34.12) |
|  | Yes | 46.97 (43.85-50.09) | 72.44 (69.65-75.23) | 77.62 (75.01-80.23) | 75.24 (72.54-77.94) | 70.01 (67.15-72.88) | 71.18 (68.36-74) | 67.55 (64.61-70.48) | 72.99 (70.21-75.77) | 73.56 (70.8-76.32) | 72.38 (69.59-75.17) | 8.76 (7-10.53) | 19.88 (17.37-22.4) | 40.83 (37.76-43.9) | 30.52 (27.65-33.39) |
| Cancer | No | 47.94 (47.28-48.6) | 70.87 (70.26-71.47) | 76.92 (76.35-77.48) | 72.33 (71.74-72.93) | 66.88 (66.26-67.51) | 68.81 (68.2-69.43) | 64.83 (64.2-65.47) | 71.19 (70.59-71.79) | 72.24 (71.64-72.83) | 68.98 (68.37-69.6) | 10.73 (10.31-11.14) | 20.17 (19.64-20.71) | 35.85 (35.22-36.49) | 33.25 (32.63-33.87) |
|  | Yes | 58.18 (52.26-64.1) | 81.52 (76.86-86.18) | 83.03 (78.53-87.53) | 82.17 (77.56-86.78) | 76.07 (70.96-81.19) | 79.19 (74.31-84.07) | 74.07 (68.87-79.28) | 76.06 (70.91-81.21) | 74.45 (69.16-79.75) | 75.4 (70.21-80.58) | 4.73 (2.29-7.18) | 17.64 (12.93-22.34) | 35.25 (29.54-40.96) | 42.38 (36.45-48.32) |
| Comorbidities | 0 | 49.16 (48.33-50) | 68.52 (67.74-69.3) | 75.82 (75.1-76.54) | 69.56 (68.79-70.33) | 65.09 (64.29-65.88) | 67.3 (66.51-68.08) | 63.25 (62.44-64.05) | 68.93 (68.16-69.71) | 71.11 (70.35-71.87) | 67.07 (66.28-67.86) | 11.94 (11.4-12.49) | 21.04 (20.36-21.73) | 33.2 (32.41-33.98) | 33.82 (33.03-34.61) |
|  | 1 | 45.47 (44.05-46.89) | 72.76 (71.48-74.04) | 77.75 (76.55-78.95) | 75.11 (73.86-76.35) | 67.84 (66.49-69.18) | 70.17 (68.85-71.49) | 65.57 (64.2-66.93) | 72.36 (71.08-73.64) | 72.72 (71.45-73.99) | 70.45 (69.14-71.76) | 9.52 (8.68-10.36) | 20.13 (18.97-21.3) | 38.4 (37.02-39.79) | 31.95 (30.62-33.27) |
|  | 2 or more | 47.24 (45.63-48.85) | 77.9 (76.56-79.24) | 80.42 (79.14-81.71) | 79.78 (78.48-81.08) | 72.96 (71.52-74.4) | 73.49 (72.07-74.91) | 70.43 (68.96-71.9) | 78.41 (77.09-79.74) | 75.95 (74.57-77.33) | 74.72 (73.31-76.12) | 7.3 (6.47-8.13) | 16.82 (15.61-18.04) | 42.49 (40.89-44.08) | 33.39 (31.87-34.91) |

Data are presented as weighted percentages with 95% confidence intervals.

### Association of Lifestyle Recommendations with Behavior Outcomes

Except for the recommendation to reduce salt intake, the delivery of lifestyle recommendations was associated with positive behavioral status (Table 4). Among current tobacco users, those who received tobacco cessation recommendation were 1.83 times (95% CI: 1.49-2.24) more likely to attempt quitting tobacco. Similarly, receiving recommendations related to physical activity and weight loss was associated with more favorable behavioral outcomes. Individuals who received recommendations to exercise and to lose weight/maintain normal weight were 1.21 times (1.13-1.29) and 1.15 times (1.07-1.22) more likely to engage in sufficient levels of physical activity, respectively. Participants who received 3-5 nutritional recommendations were 1.12 (1.04-1.21) times more likely to have a better nutritional score compared to those who received 0-2 recommendations. Those who received all six nutritional recommendations had 1.32 times higher odds (1.24-1.41) of having a better nutritional score.

**Table 4.**
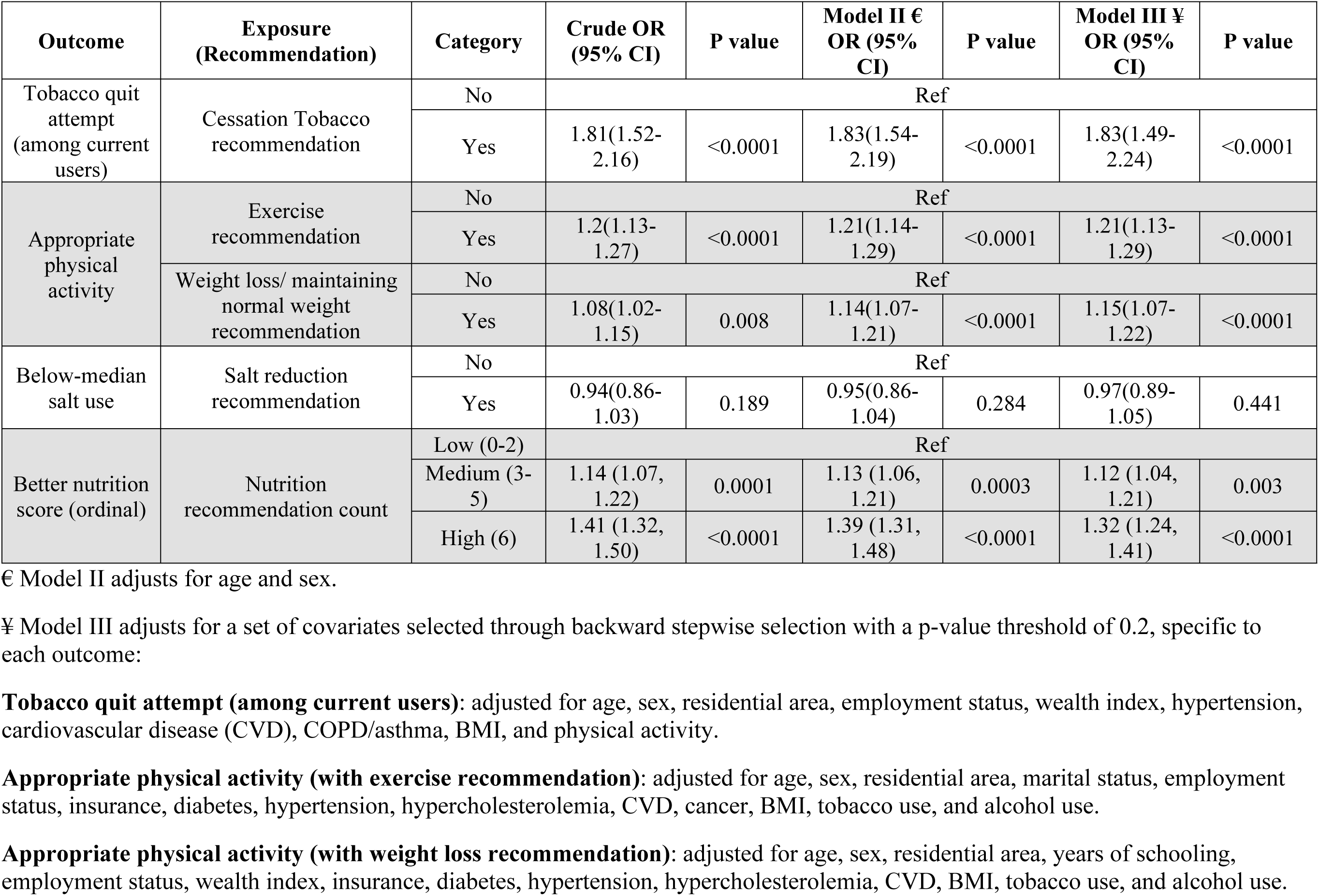
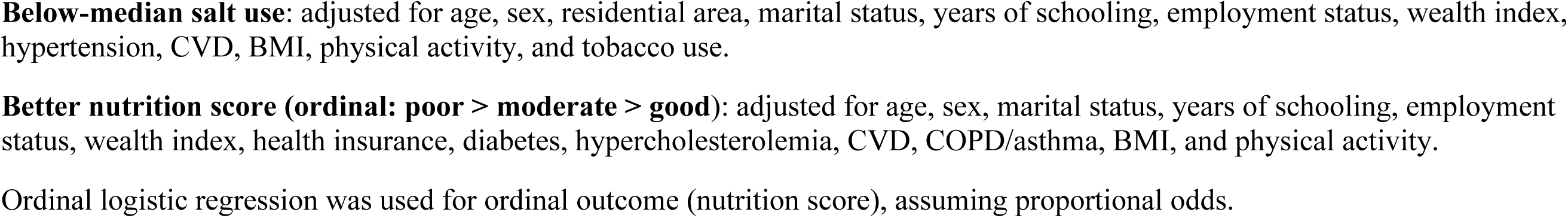
Associations Between Lifestyle Recommendations and Behavioral or Health Outcomes.

## Discussion

This study showed disparities in the delivery of lifestyle recommendations across sociodemographic groups, comorbidity, and health behavior status. Our results demonstrated that the more comorbidities an individual owns, the prevalence of lifestyle recommendation received escalated. The lifestyle recommendations delivered were shown to be associated with favorable behavioral outcomes. Notably, a dose-response relationship was observed between the number of nutritional recommendations received and improvements in dietary quality. Those individuals receiving all nutritional recommendations were 1.32 times likely to score better at nutrition scoring. Our analysis also presented a geographic variation in recommendation delivery, with the lowest coverage in eastern provinces and the highest in the northwest provinces of Iran.

In our study, lifestyle recommendations were generally provided more often to females than to males, with the exception of tobacco non-initiation/cessation advice, which was more frequently delivered to men. Pregnancy is a period during which women commonly receive lifestyle counseling, given the well-established effects of lifestyle risk factors on pregnancy outcomes[20,21], and the growing emphasis on promoting healthy behaviors during this life stage[22,23]. women tend to be more proactive in seeking healthcare and discussing health concerns, resulting in greater exposure to preventive advice. Studies show that female patients report more physical symptoms and illness behavior[24]. utilize primary care services more often for preventive and diagnostic purposes, and consult providers more frequently for both physical and mental health concerns[25,26] This greater engagement likely increases their opportunities to receive lifestyle recommendations.

Moreover, our observation that individuals with multiple comorbidities receive more health advice may be attributed to the increased frequency of healthcare visits among patients with multiple chronic conditions, providing more opportunities for counseling interactions. A 2020 study conducted across six middle-income countries found that individuals with multiple chronic conditions are more likely to have their health issues accurately identified; however, this does not necessarily lead to improved disease management or control. [27]

Our findings reinforce the role of healthcare providers in promoting healthy behaviors. Consistent with global evidence, lifestyle counseling in our study was associated with measurable behavioral gains. Participants who received weight-control or physical-activity advice were more likely to engage in sufficient activity (aOR 1.15 [1.07-1.22] and 1.21 [1.13-1.29], respectively), and smokers counseled to quit were almost twice as likely to attempt cessation (aOR 1.83 [1.49-2.24]). A graded relationship was also observed between the number of nutritional recommendations received and dietary quality (aOR 1.32 [1.24-1.41] for 6 vs ≤ 2 recommendations), echoing the dose-response patterns reported by Patnode et al. (2017) in their JAMA systematic review of behavioral counseling. Quantitatively, our effect sizes are modest but directionally consistent with U.S. NHANES analyses: Williams et al. (2021)[28] found that adults with hypertension or diabetes who received lifestyle advice had higher odds of losing weight (aOR 1.93 [1.51-2.48]), increasing physical activity (2.02 [1.73-2.37]), and improving diet (e.g., reducing sodium 4.95 [3.93-6.25]). Similarly, Davis-Ajami et al. (2021)[29] reported that prediabetic adults advised by providers were more likely to increase exercise (1.63 [1.12-2.38]) and modify diet (3.00 [1.82-4.96]). Although our population-level odds ratios are smaller, likely reflecting self-reported advice recall and single-time-point measurement, the consistency of direction supports the behavioral relevance of provider counseling. Moreover, causal evidence from an Iranian randomized trial implementing the WHO-PEN 5As model (Amini et al. 2023)[30] demonstrated significant improvements in nutrition and activity behaviors (P < 0.001) and reductions in BMI (P < 0.001), corroborating that structured, skill-based counseling can translate the associations observed in our cross-sectional data into meaningful health outcomes.

The effectiveness of lifestyle counseling depends not only on its frequency but also on how well it is targeted to individuals’ specific risk profiles. Evidence from clinical and population studies indicates that counseling yields greater benefits when directed toward higher-risk groups. For instance, the U.S. Preventive Services Task Force recommends individualized decisions about lifestyle counseling for adults without cardiovascular risk factors because benefits are modest in low-risk populations, whereas stronger evidence supports counseling for adults with obesity or cardiometabolic risk factors.[31–33] Likewise, risk-factor-targeted programs such as the Finnish post-stroke counseling model[34] and the Canadian SeeKD initiative for chronic-kidney-disease prevention show that individualized goal-oriented counseling can improve adherence, weight control, and smoking cessation among at-risk adults.[35]

Our findings are consistent with this pattern. Participants who received behavior-specific recommendations were more likely to report healthier behaviors (table 4), for example, current tobacco users advised to quit were almost twice as likely to attempt cessation (aOR 1.83, 95% CI: 1.49-2.24), and those counseled on weight control or physical activity had greater odds of meeting activity guidelines. Several other results in our study also reflected appropriate risk-based targeting. Across BMI categories, lifestyle recommendations such as reducing fat, sugar, and salt intake and increasing physical activity were more frequently provided to individuals with higher BMI, showing a consistent increase from normal weight to overweight and obese groups. Likewise, participants with a greater number of comorbidities were more likely to receive most types of lifestyle advice, and those with diabetes, hypertension, hypercholesterolemia, or cardiovascular disease were counseled more often than their healthier counterparts. These trends suggest that healthcare providers partially tailor preventive counseling to individuals’ clinical risk profiles, particularly for obesity and chronic metabolic conditions. However, our findings also revealed important deviations from this ideal pattern. For instance, tobacco counseling remained the least frequently delivered recommendation across all sociodemographic and clinical groups, even among those with multiple comorbidities or higher behavioral risk. Moreover, current smokers and alcohol consumers, despite being high-risk groups, received fewer other lifestyle recommendations. Similarly, while individuals with insufficient physical activity were more likely to be advised to exercise (74.1%, 95% CI: 73.2-75.0), they did not receive other types of lifestyle counseling more frequently, despite being at elevated overall risk. This selective emphasis indicates missed opportunities to reinforce broader preventive behaviors among high-risk individuals. Strengthening the quality and precision of targeting—so that counseling consistently reflects each patient’s risk profile—could substantially enhance the overall effectiveness of preventive care.

When comparing the provincial distribution of lifestyle counseling in this study with the spatial distribution of NCD risk factors and mortality reported by Farzadfar et al.[36] (2024), clear mismatches emerge. When comparing provincial counseling coverage with the quantitative distribution of risk factors reported by Farzadfar et al. (2024), provinces such as Sistan-Baluchestan (salt ≈ 9.2 g/day, obesity ≈ 48 % women), Hormozgan (obesity ≈ 51 %, diabetes ≈ 10 %), and Khuzestan (hypertension ≈ 40 %, diabetes ≈ 11 %), which represent some of the highest metabolic risk levels in the country, had the lowest proportions of adults receiving all ten lifestyle recommendations (< 30 %). In contrast, north-western provinces (Kurdistan, Azerbaijan, Ardabil), where smoking exceeds 40 % in men and hypertension > 40 %, demonstrated the highest counseling coverage (> 45 %), suggesting regional inequities in preventive service distribution.

The observed regional disparities in lifestyle counseling suggest a substantial unmet need for preventive advice. Such inequities reflect persistent gaps in the capacity and distribution of the preventive health workforce across Iran’s provinces. Evidence indicates that primary care physicians in rural and underserved areas often face limited training opportunities in behavioral counseling, as well as time and resource constraints.[37,38] Strengthening capacity building for health personnel—especially among physicians deployed under Iran’s mandatory rural service program—could therefore be a strategic priority. The use of virtual and blended e-learning platforms has proven effective in improving provider competencies, and can be implemented for lifestyle and NCD counseling in primary care settings. Integrating such digital training modules into existing programs like IraPEN and aligning them with WHO’s Package of Essential Noncommunicable Disease Interventions (PEN)[39] could enhance the reach and consistency of lifestyle counseling across provinces. Building a standardized, continuing education framework for preventive counseling within Iran’s PHC network would ensure that all healthcare providers, particularly those entering high-turnover rural posts, receive the skills, tools, and support needed to address patients’ behavioral risks effectively. This approach would help reduce unmet need for lifestyle advice and strengthen progress toward equitable, risk-based NCD prevention nationwide.

A major strength of this study is the use of a large, nationally and subnationally representative dataset, allowing for generalizability of findings across Iran. The comprehensive assessment of various lifestyle recommendations and their associations with both behavior and sociodemographic factors provides understanding of preventive care delivery. Moreover, the application of multivariable regression models helps control for potential confounding factors.

However, this study also has limitations. The cross-sectional design limits causal inference, as temporality between receiving recommendations and behavior change cannot be established. Additionally, all behavioral data were self-reported, which may be subject to recall or social desirability bias. Finally, the study does not capture the quality or content of lifestyle recommendations, nor the context in which they were delivered, factors that could significantly influence their effectiveness.

## Conclusion

This study provides nationally representative evidence on the delivery and effectiveness of lifestyle recommendations by healthcare providers in Iran, revealing both strengths and gaps in the current system of preventive care. Our findings demonstrate that lifestyle advice—particularly regarding diet, physical activity, and weight management—is positively associated with healthier behaviors, with a notable dose-response relationship between the number of nutritional recommendations and diet quality. The pattern of increased advice among individuals with multiple comorbidities suggests a risk-based approach by providers, yet gaps remain—especially in the under-delivery of tobacco non-initiation/cessation counseling to high-risk individuals. Geographic disparities further highlight the unequal reach of health education and intervention across provinces. These insights underscore the critical role of Iran’s healthcare system in addressing the burden of NCDs and support the integration of structured, evidence-based lifestyle counseling into routine care. Strengthening provider training, ensuring consistency in recommendation delivery, and tailoring strategies to underserved regions will be essential for advancing the country’s national NCD prevention goals and improving long-term health outcomes.

## Data Availability

The data underlying the results presented in this study are from the WHO STEPwise approach to NCD risk factor surveillance (STEPS) 2021 survey conducted in Iran. The data are owned by the Ministry of Health and Medical Education of Iran and are not publicly available. Data are available upon reasonable request from the Endocrine & Metabolism Research Institute (EMRI), Tehran University of Medical Sciences, for researchers who meet the criteria for access to confidential data. The authors did not have special access privileges.

## Acknowledgements

We would like to express our sincere gratitude to the Non-Communicable Diseases Research Center (NCDRC) and the Endocrinology and Metabolism Research Institute (EMRI) of Tehran University of Medical Sciences for their support in providing access to the 2021 WHO STEPS dataset and facilitating this research. We also extend our appreciation to the National Institute of Health Research (NIHR) for its contributions to the design, implementation, and stewardship of the STEPS survey in Iran.

We acknowledge the use of AI-assisted tools, including ChatGPT (OpenAI), for support in language editing, technical clarification, and improving the clarity of the manuscript. All intellectual content, analysis, and interpretation were performed by the authors.

## Supporting Information

**S1 Table. Scores of different categories of nutritional variables.**

